# Explainable Machine Learning models for Rapid Risk Stratification in the Emergency Department: A multi-center study

**DOI:** 10.1101/2020.11.25.20238386

**Authors:** William P.T.M. van Doorn, Floris Helmich, Paul M.E.L. van Dam, Leo H.J. Jacobs, Patricia M. Stassen, Otto Bekers, Steven J.R. Meex

## Abstract

**Background:** Risk stratification of patients presenting to the emergency department (ED) is important for appropriate triage. Diagnostic laboratory tests are an essential part of the work-up and risk stratification of these patients. Using machine learning, the prognostic power and clinical value of these tests can be amplified greatly. In this study, we applied machine learning to develop an accurate and explainable clinical decision support tool model that predicts the likelihood of 31-day mortality in ED patients (the RISK^INDEX^). This tool was developed and evaluated in four Dutch hospitals.

**Methods:** Machine learning models included patient characteristics and available laboratory data collected within the first two hours after ED presentation, and were trained using five years of data from consecutive ED patients from the Maastricht University Medical Centre+ (Maastricht), Meander Medical Center (Amersfoort), and Zuyderland (Sittard and Heerlen). A sixth year of data was used to evaluate the models using area-under-the-receiver-operating-characteristic curve (AUROC) and calibration curves. The SHapley Additive exPlanations (SHAP) algorithm was used to obtain explainable machine learning models.

**Results:** The present study included 266,327 patients with 7.1 million laboratory results available. Models show high diagnostic performance with AUROCs of 0.94,0.98,0.88, and 0.90 for Maastricht, Amersfoort, Sittard and Heerlen, respectively. The SHAP algorithm was utilized to visualize patient characteristics and laboratory data patterns that underlie individual RISK^INDEX^ predictions.

**Conclusions:** Our clinical decision support tool has excellent diagnostic performance in predicting 31-day mortality in ED patients. Follow-up studies will assess whether implementation of these algorithm can improve clinically relevant endpoints.

## Introduction

An increasing number of patients are referred to emergency departments (ED) worldwide (1, 2). Prolonged waiting times and associated crowding in the ED increase mortality, (3) and rapid risk stratification is therefore a core task in emergency medicine. An effective means to identify patients at high- and low-risk shortly after admission could help decision-making regarding patient prioritization, treatment, level of observation, and post-discharge follow-up. Consequently, numerous clinical risk scores and triage systems for stratification of patients in the ED have been developed, such as the modified early warning score (MEWS), rapid emergency medicine score (REMS) and emergency severity index (ESI) (4-7). Unfortunately, these systems often generalize poorly and lack precision, impeding their clinical use (8).

Emergency departments generate vast amounts of clinical, physical and laboratory data. This data is generally heterogeneous and comprises both structured and unstructured information. Machine learning allows processing and modelling of this data to a human interpretable level in relation to clinically relevant endpoints. Accordingly, machine learning based mortality prediction models were developed using data extracted from patients in the ED (9-13). Although these models were superior to traditional risk scores and the estimates made by physicians (9, 11, 14, 15), most are perceived as so-called “black boxes”, possibly limiting their acceptance among clinicians and raising legal or ethical concerns. Recently, models that are transparent in their patient-specific risk predictions have emerged (16-18). This development may not only advance our understanding machine learning based algorithms, but also contribute to more widespread acceptance of clinical decision-support tools based on machine learning technology among clinicians.

In this study, our aim was to develop an accurate clinical decision support tool in four hospitals in The Netherlands using machine learning technology. These models were designed to combine patient characteristics and early available laboratory results at the ED to generate an individuals’ likelihood of 31-day mortality: the RISK^INDEX^.

## Methods

### Study design and setting

A multi-center, retrospective cohort study was performed among all patients who presented to the ED at the Maastricht University Medical Center (Maastricht, The Netherlands), Meander Medical Center (Amersfoort, The Netherlands) and Zuyderland medical Center locations Sittard (Sittard, The Netherlands) and Heerlen (Heerlen, The Netherlands) between January 1, 2013 and December 31, 2018. For convenience, each of the centers will be referred to by their respective location; Maastricht, Amersfoort, Sittard and Heerlen. This study was approved by the medical ethical committees of each of the individual centers (Maastricht: #2018-0838, Amersfoort: TWO19-46, Sittard: #2018-0838, Heerlen: #2018-0838). The study follows the STROBE guidelines (19) and was conducted according to the principles of the Declaration of Helsinki (20).

### Patient population

All patients presenting to the ED aged ≥18 years with at least 3 laboratory tests ordered by the attending physician were included. Patients whose previous presentation to the ED was less than 48 hours ago were excluded.

### Dataset construction

Data anonymization, collection, processing, model selection, development and evaluation were performed for each of the four hospitals separately. All available laboratory data of the patients ordered within two hours after the first laboratory request from the ED were collected. All laboratory data acquired after two hours were not used for model development. In addition, rare laboratory tests (<0.01%) were excluded. By restricting the parameters in the model to laboratory tests ordered within the first two hours after the first test, we aimed to develop a decision support tool that would allow rapid triage after presentation. The primary outcome measure for the study was mortality within 31 days after initial ED presentation and was acquired through electronic health records.

For each hospital six years of data from consecutive patients were available. Data from the first five years was used for model development, and data from the sixth year was used to validate performance of each model. The model development data was randomly split into training (70%), tuning (20%) and calibration datasets (10%) in such a way that data from a given presentation was present in one split only.

Consequently, the training split was used to train the proposed models, the tuning set was used to iteratively improve the models by selecting the best model architectures and hyperparameters, and the calibration split was used to perform post-hoc calibration on the model predictions. Finally, the validation dataset (consisting of year six data), was used to evaluate the performance of machine learning models.

### Model selection, training and calibration

The clinical decision support tool combines patient characteristics and laboratory results through machine learning to predict the likelihood of 31-day mortality. The output of the clinical decision support tool -termed the RISK^INDEX^- is a calibrated value between 0 and 100. Various statistical and machine learning algorithms can be applied to develop such a clinical decision support tool, including regression techniques (21), neural network architectures (22), gradient boosting systems (23-25) and decision trees (26) (Supplemental section A).

The light gradient boosting system (LightGBM) architecture was selected amongst several alternatives on the basis of the tuning set performance (Supplemental information section A and Table 1). LightGBM is an implementation of distributed, efficient gradient-boosting systems with native support for missing values (25).

**Table 1.** Baseline patient and laboratory characteristics of the four study populations.

|  | **MUMC,**<br>**Maastricht**<br>N = 66,770 | **Meander,**<br>**Amersfoort**<br>N = 81,152 | **Zuyderland,**<br>**Sittard**<br>N = 66,423 | **Zuyderland,**<br>**Heerlen**<br>N = 51,982 |
| --- | --- | --- | --- | --- |
| **Demographics** |  |  |  |  |
| Age, years | 59 (± 22.2) | 59 (± 23.7) | 65 (± 21.3) | 64 (± 21.5) |
| Sex, female (%) | 32,829 (49.2%) | 41,907 (51.6%) | 34,256 (51.6%) | 26,185 (50.3%) |
| **Laboratory** |  |  |  |  |
| Mean number of tests per patient | 22 (± 10.0) | 25 (± 8.0) | 31 (± 10.4) | 31 (± 10.8) |
| Ten most frequent laboratory orders, n (%) | Creatinine, 59,937 (89.9%)<br>CBC<sup>a</sup>, 59,632 (89.4%)<br>Sodium, 56,529 (84.8%)<br>Potassium, 56,487 (84.7%)<br>CRP<sup>b</sup>, 55,178 (82.7%)<br>Urea, 53,972 (80.9%)<br>Platelets, 47,059 (70.6%)<br>Glucose, 43,733 (65.6%)<br>ALAT<sup>c</sup>, 36,429 (54.6%)<br>ASAT<sup>d</sup>, 32,130 (48.1%) | CBC<sup>a</sup>, 77,625 (95.7%)<br>CRP<sup>b</sup>, 76,639 (94.4%)<br>Sodium, 75,638 (93.2%)<br>Creatinin, 75,393 (92.9%)<br>Potassium, 75,025 (92.4%)<br>Glucose, 75,018 (92.4%)<br>Urea, 72,115 (88.9%)<br>CK, 64,356 (79.3%)<br>Platelets, 54,380 (67.0%)<br>ALAT<sup>c</sup>, 54,279 (66.9%) | CBC<sup>a</sup>, 62,518 (94.1%)<br>Platelets, 62,517 (94.1%)<br>CRP<sup>b</sup>, 60,910 (91.7%)<br>Creatinin, 60,046 (90.4%)<br>Sodium, 58,558 (88.1%)<br>Potassium, 58,404 (87.9%)<br>Glucose, 57,987 (87.3%)<br>Urea, 57,559 (86.6%)<br>ALAT<sup>c</sup>, 53,968 (81.2%)<br>ASAT<sup>d</sup>, 53,914 (81.2%) | CBC<sup>a</sup>, 49,056 (94.4%)<br>Platelets, 49,040 (94.3%)<br>CRP<sup>c</sup>, 47,573 (91.5%)<br>Creatinin, 47,043 (90.5%)<br>Sodium, 46,615 (89.7%)<br>Potassium, 46,364 (89.2%)<br>Glucose, 45,536 (87.6%)<br>Urea, 45,074 (86.7%)<br>ALAT<sup>c</sup>, 42,486 (81.7%)<br>ASAT<sup>d</sup>, 42,415 (81.6%) |
| **Outcome** |  |  |  |  |
| 31-day mortality | 4,242 (6.4%) | 3,277 (4.0%) | 3,917 (5.9%) | 2,603 (5.0%) |

<sup>a</sup> Complete Blood Count including hemoglobin, hematocrit, MCH, MCV and white blood cells; <sup>b</sup> C-reactive Protein; <sup>c</sup> Alanine (Amino)Transaminase; <sup>d</sup> Aspartate (Amino)Transaminase

Consequently, a broad spectrum of hyperparameter combinations for this architecture was evaluated (Supplemental Table 2). Hyperparameter optimization is the process of selecting a set of optimal hyperparameters, which are features controlling the training process of a machine learning model; such as the rate of learning and the maximum level of complexity. In the current study, bayesian hyperparameter optimization using tree-parzen estimators (TPE) was utilized (27). This approach is based on building a probability model of the objective function and using this to select the most promising hyperparameters to evaluate in the true objective function. Optimization was run for 1,000 iterations with logarithmic loss as our objective function (Supplemental Table 2 lists the definitions of the search space per hyperparameter). Hyperparameter optimization resulted in LightGBM architectures consisting of 220 – 740 boosted trees with a maximum depth of 11 – 37 and maximum leaves of 320 – 690 for each base learner (see Supplemental Table 3). Exponential learning-rate decay was used during training with an initial learning rate of 0.075 – 0.145 decaying every 2,000 training steps by a factor of 0.7-0.8. The loss function during training was logarithmic loss.

**Table 2.** Illustrative example of clinical decision support tool using developed machine learning models. The fixed-threshold decision support tool was fixed at a negative predictive value of 99% to identify low-risk patients (corresponding RISK^INDEX^ cut-offs between 1.0 and 12.1) and fixed at a positive predictive value of 75% to identify high-risk patients (corresponding RISK^INDEX^ cut-offs between 29.9 and 64). Diagnostic metrics and proportion of patients identified as either low- or high-risk are described in the table.

| **Low-risk**<br>defined at a negative predictive value of 99% |  |  |  |  |  |
| --- | --- | --- | --- | --- | --- |
|  | **RISK<sup>INDEX</sup>**<br>**cut-off** | **Sensitivity** | **Specificity** | **NPV** | **Proportion**<br>**of patients** |
| Maastricht | 4.3 | 86.9%<br>[84.2% - 88.9%] | 87.7%<br>[87.2% - 88.2%] | 99.0%<br>[98.8% - 99.2%] | 83.0%<br>[82.3% - 83.6%] |
| Meander | 12.1 | 75.9%<br>[72.3% - 78.6%] | 97.5%<br>[97.3% - 97.7%] | 99.0%<br>[98.8% - 99.1%] | 94.7%<br>[94.3% - 95.0%] |
| Sittard | 1.0 | 85.5%<br>[83.2% - 87.3%] | 75.0%<br>[74.2% - 75.7%] | 98.8%<br>[98.6% - 99.0%] | 71.5%<br>[70.7% - 72.0%] |
| Heerlen | 1.0 | 82.7%<br>[80.0% - 85.2%] | 81.2%<br>[80.8% - 81.8%] | 98.9%<br>[98.8% - 99.1%] | 78.1%<br>[77.7% - 78.8%] |
| **High-risk**<br>defined at positive predictive value of 75% |  |  |  |  |  |
|  | **RISK<sup>INDEX</sup>**<br>**cut-off** | **Sensitivity** | **Specificity** | **PPV** | **Proportion**<br>**of patients** |
| Maastricht | 41.1 | 45.7%<br>[42.3% - 49.5%] | 99.0%<br>[98.8% - 99.1%] | 74.9%<br>[70.8% - 77.8%] | 3.9%<br>[3.5% - 4.2%] |
| Meander | 29.9 | 60.6%<br>[56.9% - 64.1%] | 99.2%<br>[99.0% - 99.3%] | 74.9%<br>[70.8% - 78.3%] | 3.1%<br>[2.9% - 3.4%] |
| Sittard | 64 | 14.7%<br>[12.6% - 16.7%] | 99.7%<br>[99.6% - 99.8%] | 74.8%<br>[68.8% - 80.6%] | 1.1%<br>[1.0% - 1.3%] |
| Heerlen | 41.1 | 27.1%<br>[24.2% - 29.4%] | 99.5%<br>[99.4% - 99.7%] | 74.8%<br>[70.6% - 80.6%] | 1.7%<br>[1.5% - 2.0%] |

LightGBM models with optimal hyperparameters were recalibrated on the calibration set in order to further improve the quality of the generated RISK^INDEX^. Recalibration is recommended as most LightGBM models are prone to miscalibration, meaning that their output RISK^INDEX^ do not represent the actual 31-day mortality likelihood. Hence, recalibration ensures that consistent probabilistic interpretations of the RISK^INDEX^ predictions can be made (28). For calibration, various techniques were considered including Platt scaling (29), isotonic regression (30) and Platt-Binner scaling (31). Model calibration was assessed by the brier score (32) and visual inspection of reliability plots (33). Reliability plots are the usual approach for evaluating calibration of binary outcomes in which we compare decile-binned means of predictions versus means of the observed outcomes in the patients. Platt-Binner scaling was selected as this was shown to result in the best calibrated models (see Supplemental Figure 1). The resulting calibrated predictions were defined as the RISK^INDEX^. Data preprocessing, model development, selection, training and calibration was performed using Python programming language (version 3.7.1) using packages Numpy (version 1.17), Pandas (version 0.24), Keras (version 2.2.2), scikit-learn (version 0.22.0) and tensorflow (version 2.0.1, beta).

**Figure 1.**
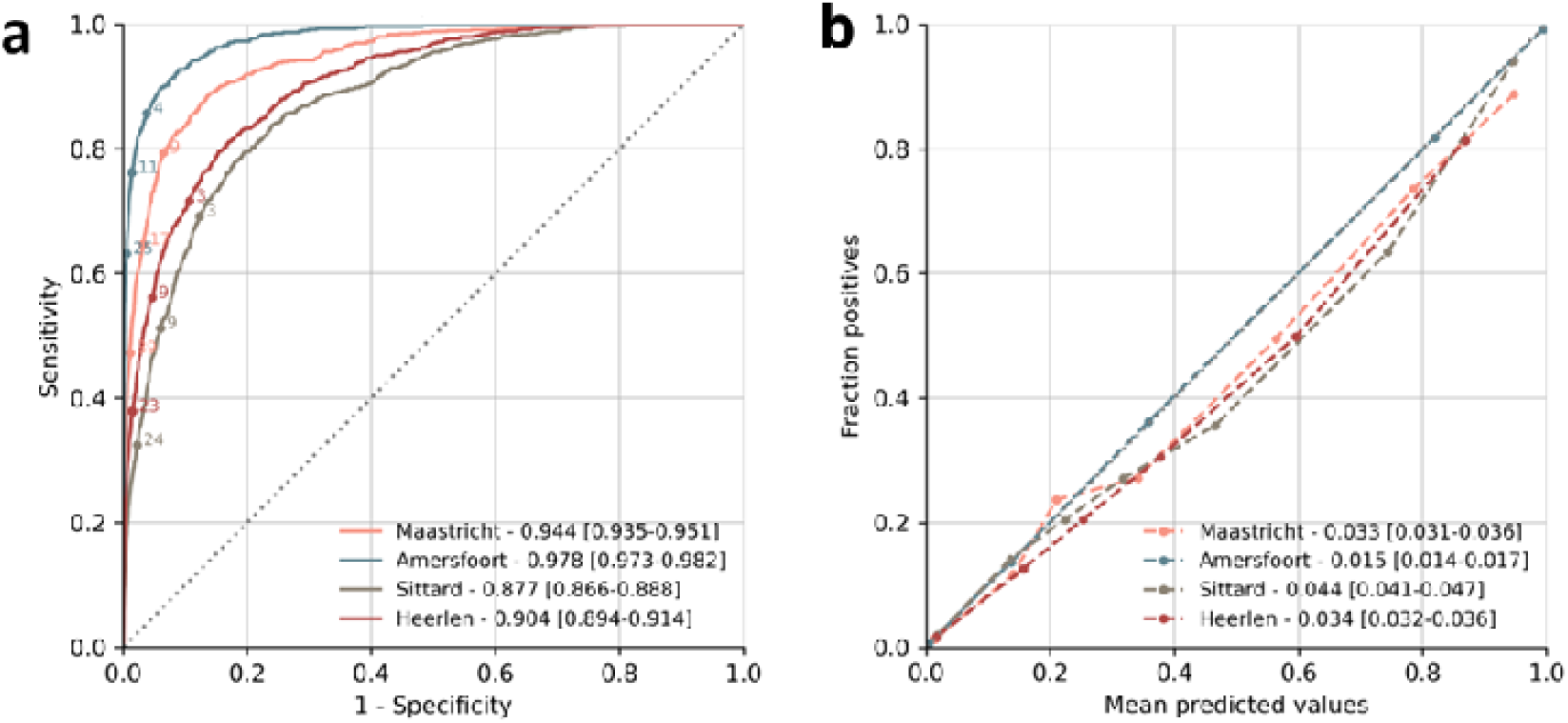
Discrimination and calibration of machine learning models. (A) Receiver operating characteristic curves (ROC) showing the discrimination of the LightGBM models in each of the different centers. Annotated points depict example RISK^INDEX^ thresholds for illustrative purposes. (B) Calibration of the machine learning models with the observed proportion of 31-day mortality in each of the centers. Each point represents 10% of the patients in the validation dataset.

### Model evaluation

Overall model performance was evaluated in the validation set of each hospital separately, all of them containing 1 full year of data from ED patients, not previously used for model development. Evaluation was done by 1) area under the receiver-operating-characteristic curve (AUROC) to quantify the ability of models to discriminate between survivors and non-survivors, and 2) visual inspection of calibration curve and brier scores to estimate how accurately the RISK^INDEX^ estimates the likelihood of 31-day mortality. Next, an embedded reference table was created based on the validation dataset to report estimates of sensitivity, negative predictive value (NPV), specificity and positive predictive value (PPV) for each RISK^INDEX^ between 0-100. This table was subsequently used to compare diagnostic metrics from the model (sensitivity, NPV, specificity, and PPV) across the four hospitals at various selected statistical thresholds.

### Model explanation

To facilitate the interpretation of the RISK^INDEX^ generated by our machine learning models, the Shapley additive explanations (SHAP) algorithm was applied (16, 34). SHAP highlights the patient characteristics and laboratory results (further referred to as “variables”) that underlie patient-specific predictions by the model, hence mitigating the issue of black-box predictions. SHAP is a model-agnostic representation of feature importance where the impact of each variable on a particular prediction is represented using Shapley values inspired by cooperative game theory and their extensions (35-37). A Shapley value states -given the current set of variables- how much a variable in the context of its interaction with other variables contributes to the difference between the actual prediction and the mean prediction. That is, the mean prediction plus the sum of the Shapley values for all variables equals the actual prediction. It is critically important to understand that this is fundamentally different to direct variable effects known from e.g. (generalized) linear models. The SHAP value for a variable should not be seen as its direct -and isolated effect- but as its aggregated effect when interacting with other variables in the model. In our specific case, positive Shapley values contribute towards a positive prediction (death), whilst low or negative Shapely values contribute towards a negative prediction (survival).

### Statistical analysis

Descriptive analysis of baseline characteristics was performed using IBM SPSS Statistics for Windows (version 24.0). Continuous variables were reported as means with standard deviation (SD) or medians with interquartile ranges (IQRs) depending on the distribution of the data. Categorical variables were reported as proportions. Thousand bootstrap iterations were used to calculate 95% confidence intervals, unless otherwise mentioned. Model evaluation and statistical analysis was performed using Python (version 3.7.1) using packages Numpy (version 1.17), Pandas (version 0.24) and Matplotlib (version 3.1.2).

## Results

### Patient and laboratory characteristics

In the current study more than 50,000 presentations were included for each hospital resulting in a total of 266,327 unique presentations to the ED. The total population consisted of slightly more female (mean; 50.8%) patients with a mean age of 61.5 (+- 22.4) years. Within the first two hours of presentations, on average 29 (± 10.6) laboratory parameters were requested. Although requested parameters were subject to intercenter heterogeneity, complete blood count, electrolytes and lactate dehydrogenase (LD) represented the most prevalent in all hospitals. Mortality rates at 31 days were 6.4%, 4.0%, 5.9% and 5.0% for Maastricht, Amersfoort, Sittard and Heerlen, respectively. Baseline characteristics are described in Table 1.

### Model performance

Using the available patient characteristics and laboratory data from the first two hours of a presentation, we developed machine learning models that predict the 31-day mortality likelihood of an individual patient presenting to the ED: the RISK^INDEX^. Machine learning models were able to discriminate between patients who died or survived within 31-days as depicted by AUROCs of 0.944 [0.935-0.951], 0.978 [0.973-0.982], 0.877 [0.866-0.888] and 0.904 [0.894-0.914] for Maastricht, Amersfoort, Sittard and Heerlen, respectively (Figure 1A). After calibration of the raw output scores (see supplemental Figure 1), the RISK^INDEX^ corresponded well with the likelihood of 31-day mortality (Figure 1B) confirmed by brier scores of 0.033 [0.031-0.036], 0.015 [0.014 – 0.017], 0.044 [0.041-0.047] and 0.034 [0.032-0.036] for Maastricht, Amersfoort, Sittard and Heerlen, respectively. Hence, the RISK^INDEX^ provides an individualized and precise assessment of 31-day mortality risk by combining patient characteristics and all available laboratory data available within the first two hours after ED presentation.

### From theoretical model to clinical decision support tool

The RISK^INDEX^ is by design a continuous measure, with a high RISK^INDEX^ translating to a high likelihood of 31-day mortality and low RISK^INDEX^ translating to low likelihood of 31-day mortality (see Supplemental Figures 1-4). We recognize, however, that in clinical practice most decision support tools use fixed thresholds to categorize patients as low-, medium- or high-risk. Consequently, our RISK^INDEX^ can readily be transformed to such a fixed-threshold decision support tool, and users can control thresholds in accordance to the desired risk tolerance level. An illustrative example of how an individual hospital may employ the RISK^INDEX^ is as follows: define the acceptable percentage of patients that are erroneously identified as “low-risk” by the model (any number from 0-100). This percentage, e.g. 1%, could be derived from an inventory of acceptable risk tolerance for adverse events by patients, health care workers, or both (38). Then, use the corresponding negative predictive value (in this case 99%) to derive the matching RISK^INDEX^ threshold from the calibration set and associated values for sensitivity, specificity, and proportion of subjects identified as low risk (Table 2). A similar approach can be applied to identify high-risk patients: define the positive predictive value that would provide an acceptable balance between true high risk patient identification and false positives, e.g. a positive predictive value of 75% would categorize between 1.1% and 3.9% as high-risk individuals with 1 in 4 “flaggings” by the clinical decision support tool being false positive (Table 2). A higher proportion of high-risk subject identification is feasible but will be at the expense of increased false positive flaggings.

### Explainable model predictions

To understand the patterns underlying the RISK^INDEX^ generated for each individual patient, the SHAP algorithm was applied (see Supplemental Figures 6-8). This is illustrated for a low-, medium and high-risk individual in Figure 2. A high RISK^INDEX^ was generated for a 71-year old (+5.5 RISK^INDEX^) individual with a high numeric amount of laboratory measurements (+31.5 RISK^INDEX^), a high lactate dehydrogenase (LD; +17.7) and a low albumin level (+6), whilst the remainder of the features contributed another significant portion (+10.6) ultimately leading to a RISK^INDEX^ of 76. On the other hand, the low-risk individual had a normal albumin level (−0.9 RISK^INDEX^), the presence of a pH blood measurement (−0.6), a normal lymphocyte level (−0.5) and the remaining variables further lowered the prediction (−1.3). Yet, a relatively high urea level (17 mmol/L) caused a small increase in RISK^INDEX^ (+0.7). These figures exemplify the explainability of our machine learning models.

**Figure 2.**
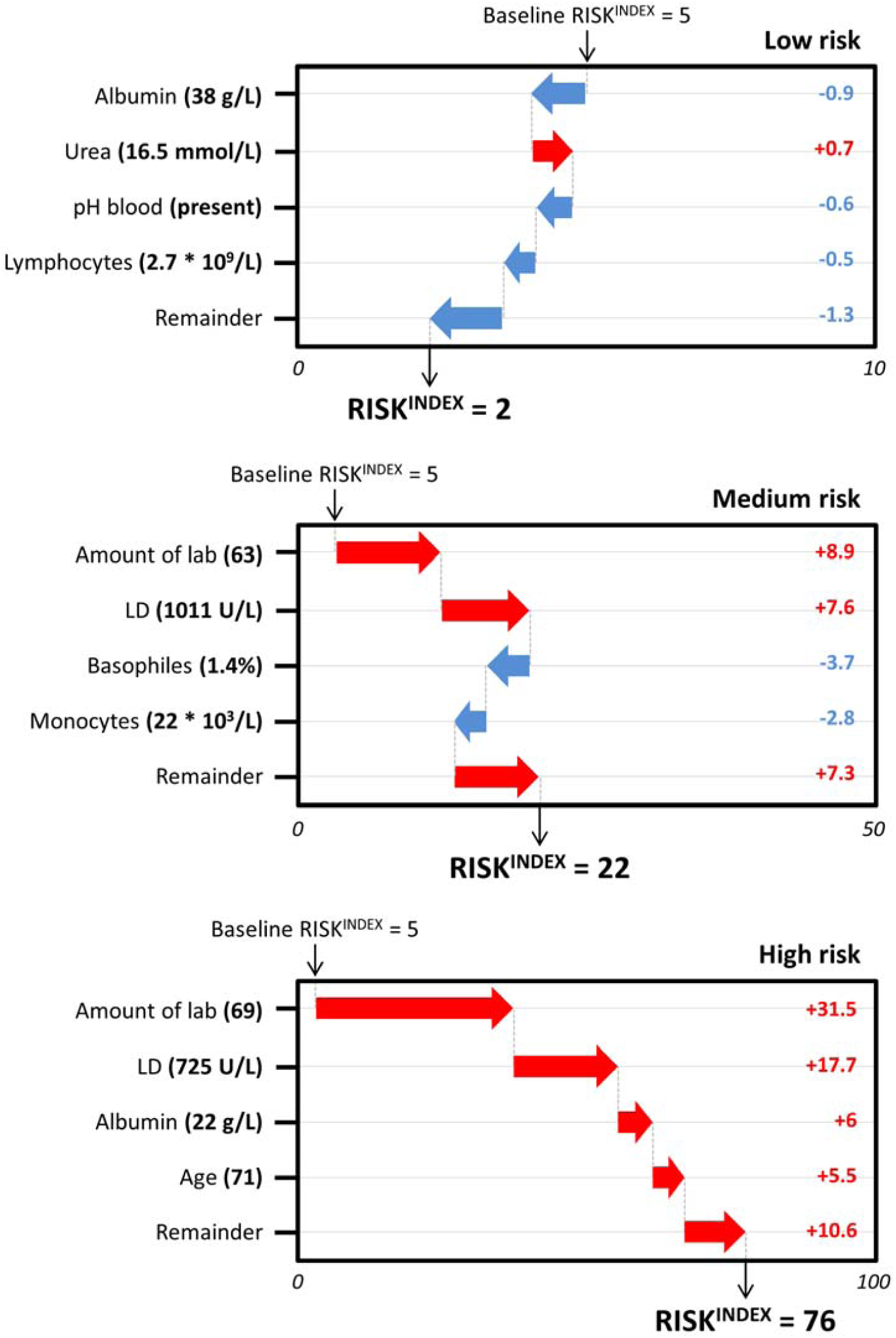
Illustrative example of how patient characteristics and laboratory results build up to a RISK^INDEX^. Illustrative example of how patient characteristics and laboratory results build up to a RISK^INDEX^ in a low-risk (upper panel, RISK^INDEX^ of 2), medium-risk (mid panel, RISK^INDEX^ of 22), and high-risk patient (lower panel, RISK^INDEX^ of 76).

## Discussion

In a large, multi-center study including over 260,000 patients presenting to the emergency department across four hospitals, machine learning technology was used to develop and evaluate novel clinical decision support tools that incorporate baseline patient characteristics and laboratory data to accurately predict the likelihood of 31-day mortality. Explainable machine learning models were utilized that were well calibrated and had overall high diagnostic performance. Our study has several unique characteristics.

First, our clinical decision support tool provides an individualized, precise, and rapid assessment of 31-day mortality risk -the RISK^INDEX^- by using patient characteristics and baseline laboratory results acquired within two hours after the presentation of the patient. Our models had high diagnostic performance with AUROCS of 0.944 [0.935-0.951], 0.978 [0.973-0.982], 0.877 [0.866-0.888] and 0.904 [0.894-0.914] (Maastricht, Amersfoort, Sittard and Heerlen), outperforming any clinical decision support tool or risk score currently used in the emergency department for risk stratification (5, 6, 39).

Second, the Shapley additive explanations (SHAP) algorithm was used to obtain explainable machine learning models (16, 17). The SHAP algorithm facilitates the interpretation of patient characteristics and laboratory results that drive patient-specific RISK^INDEX^ predictions (as illustrated in Figure 2). Development of such explainable machine learning models mitigates the issue of “black-box” predictions, and contributes to the understanding and acceptance of these models amongst clinicians and nurses. Furthermore, transparency in these models will likely become inevitable as international regulators have expressed concerns regarding black-box predictions, signaling that automated prediction systems are enforced to inform users about the logic involved, as well as the significance and the envisaged consequences of its predictions in the near future (40).

Third, our clinical decision support tool is highly versatile as it can be adjusted to the demands of each specific healthcare system or institution. For example, a triage algorithm was illustrated using a negative predictive value of 99% to identify low-risk patients, and a positive predictive value of 75% to identify high-risk patients (Table 2). Nevertheless, in case a more conservative policy would be desired, the low-risk thresholds can be adjusted accordingly, e.g. to an even higher NPV of 99.5% implying that only 5 out of a 1.000 patients would erroneously be identified as “low-risk”. Implementation of such a triage system using our proposed clinical decision tool is convenient as current models rely on data that are easily collected through existing laboratory system infrastructure. This is an advantage compared to machine learning models trained with e.g. unstructured clinical data that require manual annotation or natural language processing.

Fourth, application of our methodology to four separate hospitals provides support for its robustness and consistency. Differences in diagnostic performance between hospitals can in part be explained by demographic differences, patient mix (and hence baseline mortality rates), and the laboratory testing patterns of the attending physicians.

Last, the large sample size of more than 260,000 patients and 7.1 million laboratory tests allowed for the development of machine learning models with high performance. Despite our models being trained almost exclusively with laboratory data, they outperform machine learning models which also had full access to clinical data of a patient (12, 13). This highlights that high-performance machine learning models -besides having access to as much individual patient data-require large sample sizes in order to achieve optimal performance for a specific prediction task.

### Literature

A limited number of attempts to use machine learning technology for risk stratification in the ED in a retrospective setting has been described (9, 10, 12, 13). Klug et al. and Perng et al. developed machine learning models with performance in line with our study (AUCs of 0.96 and 0.93, respectively) (12, 13). Although diagnostic performance was similar, there are some notable differences. First, these studies focused on populations from a single center. Second, these studies used clinical and vital characteristics of patients whereas our study almost exclusively relied on the laboratory results, which makes it easier to implement and extend to other hospitals. Third, the interpretation of our generated RISK^INDEX^ was facilitated on a patient level using the recent SHAP algorithm. Fourth, illustrative implementation strategies were provided using pre-defined safety (NPV) and efficacy (PPV) measures to identify low- and high-risk patients at the emergency department, respectively.

### Limitations

Several limitations should be recognized. First, the current study is based on retrospective data and prospective studies are desired to study performance and true clinical benefit of our clinical decision support tool in a real-world setting. This would also allow us to study the (dis)advantages of implementing these models using a triage system based on statistical thresholds compared to an approach based on individual RISK^INDEX^ estimates. Second, these models possess -despite being explainable-algorithmic bias; models have been trained entirely upon the basis of what humans have done before. This implies that the predictions of our model cannot be extrapolated, and that predictions in e.g. minority populations have a higher degree of uncertainty. To facilitate the interpretation of such uncertain predictions, it would be interesting to implement uncertainty measures amongst the prediction, e.g. in form of confidence intervals. This could warn clinicians when a certain prediction is highly uncertain, potentially leading to increased trust and interpretability amongst the users of these clinical decision support tools. This is novel development in machine learning which requires additional technical developments.

## Conclusion

Our novel RISK^INDEX^ clinical decision support tool incorporates patient characteristics and laboratory tests available within the first two hours after presentation to provide an individual, precise and rapid assessment of the patient’s mortality risk within 31 days. These models had overall high diagnostic performance, are explainable, and can be implemented in a triage system extending current systems used in modern emergency departments. Prospective, follow-up studies are warranted to study the performance and clinical benefit of these models in a real-world clinical setting.

## Data Availability

All relevant data are within the manuscript and its Supporting Information files. The raw
data and analysis code for this study are available on reasonable request.

## Acknowledgements

None.

