## Supplemental for "Explainable Machine Learning models for Rapid Risk Stratification in the Emergency Department: A multi-center study"

**Supplemental Information**

Supplemental section A. Information of baseline model architectures.

**Supplemental Tables**

Supplemental Table 1. Baseline comparison of statistical and machine learning models.

Supplemental Table 2. Hyperparameter search space.

Supplemental Table 3. Hyperparameter settings for LightGBM models.

**Supplemental Figures**

Supplemental Figure 1. Model calibration.

Supplemental Figure 2. Embedded reference figures for machine learning model in Maastricht.

Supplemental Figure 3. Embedded reference figures for machine learning model in Amersfoort.

Supplemental Figure 4. Embedded reference figures for machine learning model in Sittard.

Supplemental Figure 5. Embedded reference figures for machine learning model in Heerlen.

Supplemental Figure 6. Impact of model parameters.

Supplemental Figure 7. Global SHAP values in each of the centers.

Supplemental Figure 8. Impact of individual features on the SHAP value.

**Supplemental Information**

**Supplemental section A. Information of baseline model architectures.** A comparison of available algorithms was conducted on the 31-day mortality prediction task. The following algorithms:

- **Logistic regression:** a simple logistic regression model was used with L2 regularization, a tolerance of 1^e-4^ and a maximum amount of iterations of 1,000. We used the limited Broyden–Fletcher–Goldfarb–Shanno (lbfgs) algorithm as optimizer. Logistic regression was implemented using Python (version 3.7.1) and the scikit-learn (version 0.22.1) package.
- **Feed-forward neural networks:** a simple feed-forward, multi-layer perceptron neural network was implemented consisting of three hidden layers with respectively 128, 64 and 32 neurons and RELU activation functions. We trained the network using a constant learning rate of 0.001 with a batch size of 1 and the Adam optimization scheme. The neural network was implemented using Python programming language (version 3.7.1) using packages Keras (version 2.2.2) and scikit-learn (version 0.22.1).
- **Random Forest:** a random forest classifier with a decision tree as base learner, consisting of 300 trees with gini criterion and a maximum depth of 50 was used. We used bootstrapped samples for building trees. Random forest was implemented using Python (version 3.7.1) and the scikit-learn (version 0.22.1) package
- **Gradient-boosting systems:** we used three different implementations of gradient-boosting systems: CatBoost (1), XGBoost (2) and LightGBM (3). Each of them has specific unique implementation details, but all use decision trees as the base weak learner and gradient boosting to iteratively fit a sequence of such trees. To provide a valuable baseline comparison, we aimed to evaluate these implementations with as identical hyperparameters as possible. Thus, for each implementation, we used a learning rate of 0.01, a maximum number of trees of 500 and a maximum depth of each base learner to be 50. We implemented this using the Python programming language (version 3.7.1) using packages XGBoost (version 0.90), CatBoost (version 0.20.2) and LightGBM (version 2.3.1).

**Supplemental Tables**

**Supplemental Table 1. Baseline comparison of statistical and machine learning models.** Comparison of statistical and machine learning models for the prediction of 31-day mortality in validation dataset. Models were trained on the training dataset and their performance was evaluated on the tuning dataset. Performance was assessed using their discrimination ability by ROC curves, and their calibration performance by using brier scores. 95% confidence intervals were calculated using 1,000 bootstraps.

|  | **Maastricht** | | **Sittard** | | **Heerlen** | | **Amersfoort** | |
| --- | --- | --- | --- | --- | --- | --- | --- | --- |
| **Baseline model** | AUC | Brier | AUC | Brier | AUC | Brier | AUC | Brier |
| Logistic regression | 0.73 [0.708 - 0.748] | 0.06 [0.060 - 0.068] | 0.62 [0.581 - 0.625] | 0.06 [0.055 - 0.066] | 0.67 [0.655 - 0.694] | 0.05 [0.049 - 0.054] | 0.79 [0.767 - 0.803] | 0.04 [0.034 - 0.039] |
| Neural network | 0.76 [0.740 - 0.779] | 0.06 [0.055 - 0.063] | 0.78 [0.751 - 0.812] | 0.06 [0.048 - 0.059] | 0.78 [0.759 - 0.796] | 0.04 [0.039 - 0.045] | 0.81 [0.788 - 0.823] | 0.04 [0.038 - 0.043] |
| Random Forest | 0.77 [0.752 - 0.789] | 0.05 [0.051 - 0.057] | 0.76 [0.743 - 0.777] | 0.05 [0.050 - 0.052] | 0.76 [0.738 - 0.773] | 0.04 [0.040  -0.045] | 0.80 [0.781 - 0.817] | 0.04 [0.032 - 0.041] |
| CatBoost | 0.89 [0.879 - 0.901] | 0.05 [0.042 - 0.047] | 0.86 [0.855 - 0.0873] | 0.05 [0.042 - 0.049] | 0.90 [0.898 - 0.909] | 0.03 [0.030 - 0.037] | **0.94 [0.924- 0.951]** | **0.02 [0.016 - 0.02]** |
| XGBoost | 0.87 [0.856 - 0.880] | 0.05 [0.045 - 0.051] | 0.87 [0.857 - 0.878] | 0.05 [0.043 - 0.049] | 0.87 [0.865 - 0.890] | 0.04 [0.034 - 0.040] | 0.92 [0.913 - 0.931] | 0.03 [0.024 - 0.035] |
| LightGBM | **0.91 [0.898- 0.912]** | **0.04 [0.037 -0.042]** | **0.88 [0.869 -0.889]** | **0.04 [0.041**  **-0.047]** | **0.90 [0.897- 0.911]** | **0.03 [0.031- 0.036]** | 0.93 [0.922 - 0.948] | 0.02 [0.019 - 0.028] |

**Supplemental Table 2. Hyperparameter search space.** Hyperparameter combinations evaluated in the current study to find optimal hyperparameters for LightGBM architectures.

| **Hyperparameter** | **Values considered** |
| --- | --- |
| *Data preprocessing* | |
| Normalization to [0, 1] | On, off |
| Presence of binary ‘presence’ variable | On, off |
| Presence of numeric amount variable | On, off |
| *Gradient-boosting tree architecture* |  |
| Number of boosted trees to fit | 10 - 1000 on a 10 scale |
| Maximum amount of tree leaves for base learner | 10 - 1000 on a 10 scale |
| Maximum tree depth for base learner | 1 - 50 on a 1.0 scale |
| Minimum sum of instance weight needed in a leaf | 0 - 10 on a 1.0 scale |
| Subsample ratio of columns when constructing each tree. | 0.01 to 1.0 on a 0.01 scale |
| *Model training* | |
| Learning rate | 0.025 - 1.0 on a 0.005 scale |
| Learning rate scheduling | On, off |
| Learning rate scheduling decay | 0.5, 0.6, 0.7, 0.8, 0.9, 0.95 |
| Subsample ratio of the training instance. | 0.5 to 1.0 on a 0.05 scale |
| L1 regularization term on weights. | 1^e^-9 to 1^e^3 on a log-scale |
| L2 regularization term on weights*.* | 1^e^-9 to 1^e^3 on a log-scale |

**Supplemental Table 3. Hyperparameter settings for LightGBM models.** Hyperparameter settings for each of the LightGBM models developed in Maastricht, Sittard, Heerlen and Amersfoort, respectively.

| **Hyperparameter** | **MUMC**  **Maastricht** | **Zuyderland**  **Sittard** | **Zuyderland**  **Heerlen** | **Meander**  **Amersfoort** |
| --- | --- | --- | --- | --- |
| *Data preprocessing* | | | | |
| Normalization to [0, 1] | Off | Off | Off | Off |
| Presence of binary ‘presence’ variable | On | On | On | On |
| Presence of numeric amount variable | On | On | On | On |
| *Gradient-boosting tree architecture* | | | | |
| Number of boosted trees to fit | 370 | 690 | 310 | 420 |
| Maximum amount of tree leaves for base learner | 240 | 940 | 640 | 220 |
| Maximum tree depth for base learner | 23 | 14 | 13 | 47 |
| Minimum sum of instance weight needed in a leaf | 2.0 | 7.0 | 3.0 | 4.0 |
| Subsample ratio of columns when constructing each tree | 0.809 | 0.681 | 0.971 | 0.824 |
| *Model training* | | | | |
| Learning rate | 0.075 | 0.090 | 0.135 | 0.145 |
| Learning rate scheduling | On | On | On | On |
| Learning rate scheduling decay | 0.8 | 0.8 | 0.7 | 0.8 |
| Subsample ratio of the training instance. | 0.75 | 0.70 | 0.75 | 0.65 |
| L1 regularization term on weights. | 2.435 e-9 | 4.512 e-7 | 3.743 e-7 | 2.799 e-9 |
| L2 regularization term on weights*.* | 4.593 e-6 | 5.329 e-1 | 4.529 e-2 | 1.069 e-6 |

**Supplemental Figures**

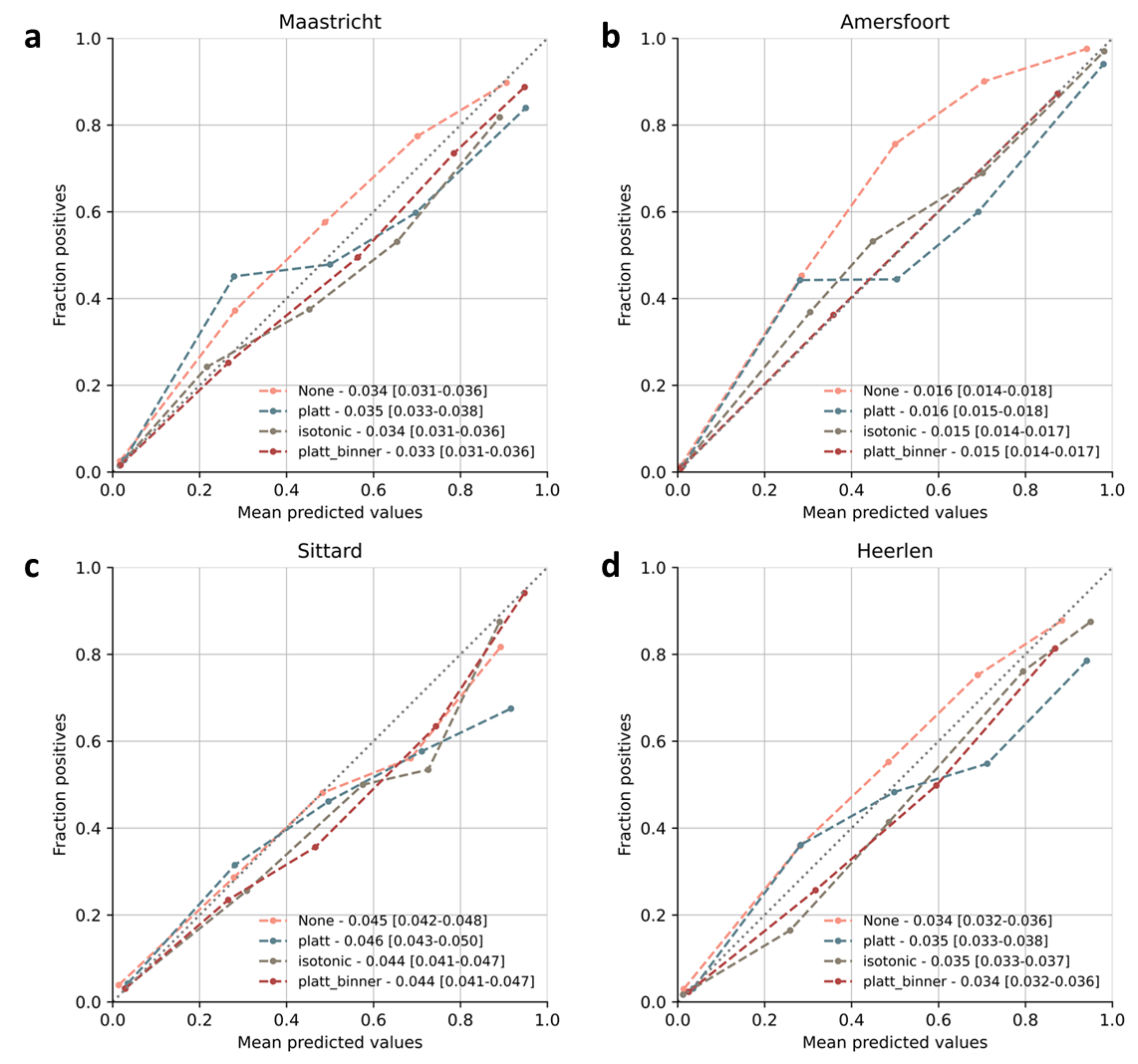

**Supplemental Figure 1. Model calibration.** To ensure that we can make probabilistic interpretations of our proposed RISK^INDEX^, we need to confirm that the predicted RISK^INDEX^ corresponds with the likelihood of 31-day mortality. As expected due to the nature of our machine learning architecture, our raw, i.e. uncalibrated, models did not provide us with well-calibrated values (see A-D). Calibration with the Platt-Binner technique, calibration curves show well calibrated algorithms confirmed by low Brier scores of 0.033 [0.031-0.036], 0.015 [0.014-0.017], 0.044 [0.041-0.047] and 0.034 [0.032-0.036] for each of the centers as previously described.

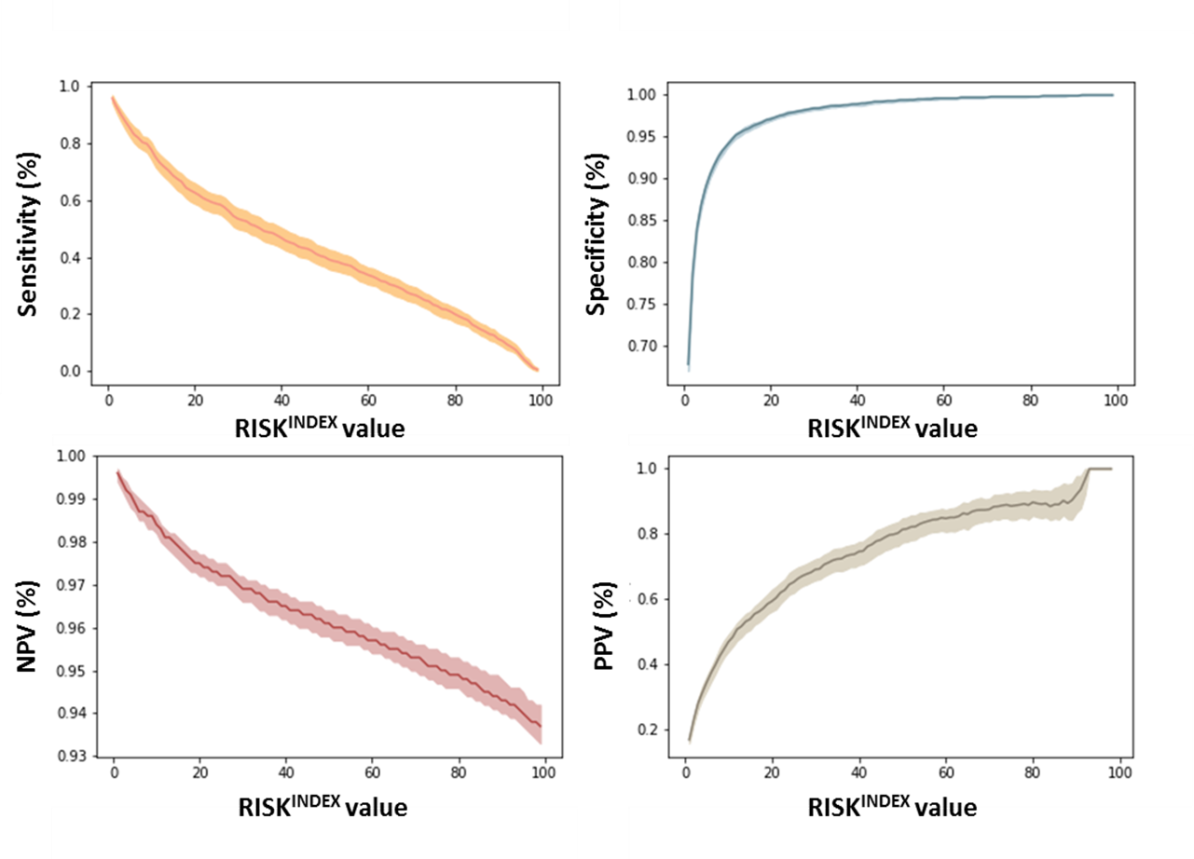

**Supplemental Figure 2. Embedded reference figures for machine learning model in Maastricht.** For each patient in the Maastricht University Medical Center the model outputs a RISK^INDEX^ value which corresponds to an estimated sensitivity, specificity, PPV and NPV with 95% confidence intervals. Lines are the point estimates at each RISK^INDEX^ and shaded regions are the 95% confidence intervals.

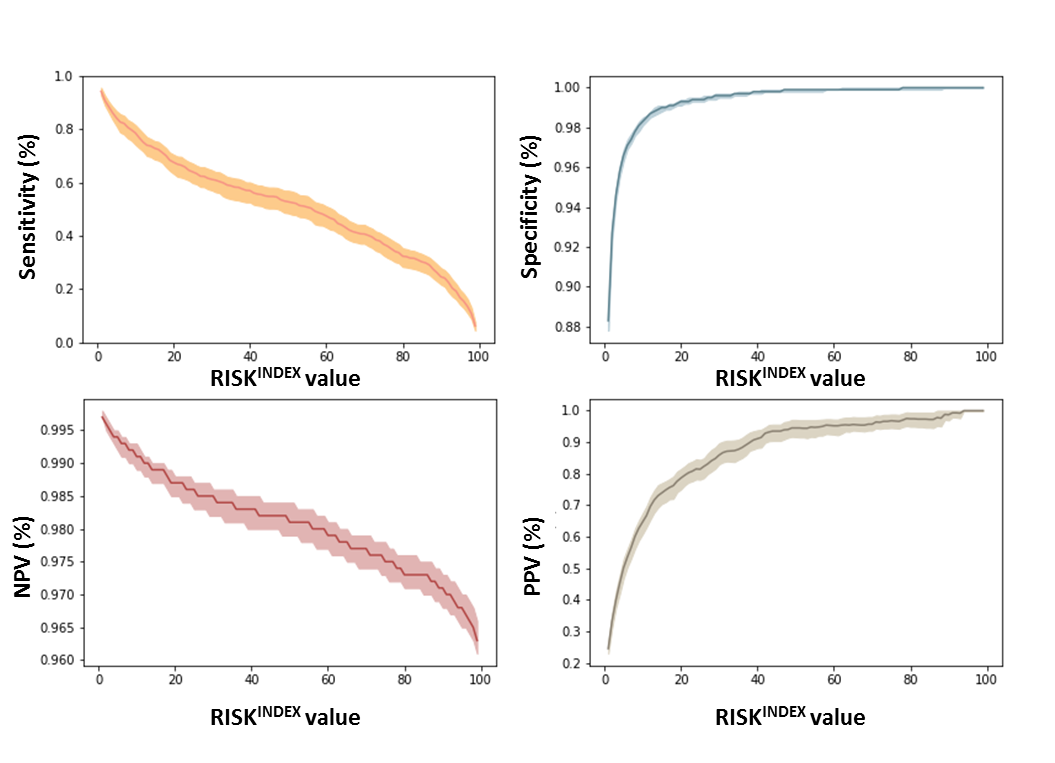

**Supplemental Figure 3. Embedded reference figures for machine learning model in Amersfoort.** For each patient in the Meander Medical Center (Amersfoort) the model outputs a RISK^INDEX^ value which corresponds to an estimated sensitivity, specificity, PPV and NPV with 95% confidence intervals. Lines are the point estimates at each RISK^INDEX^ and shaded regions are the 95% confidence intervals

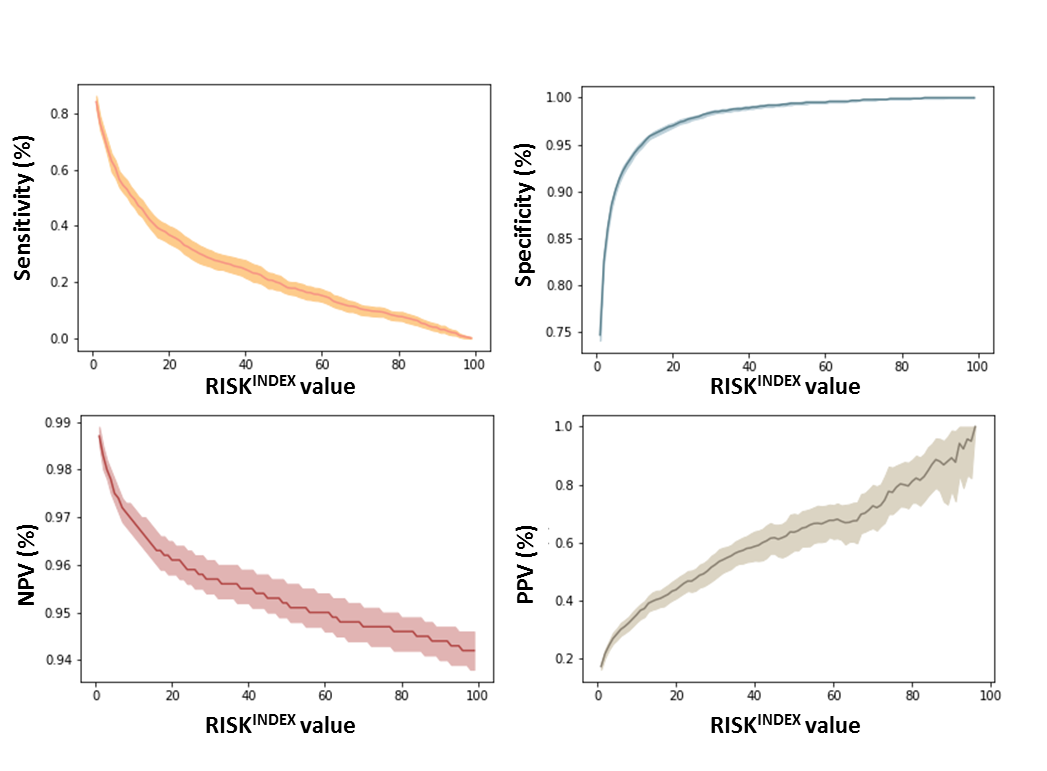

**Supplemental Figure 4. Embedded reference figures for machine learning model in Sittard.** For each patient in the Zuyderland medical center, location Sittard the model outputs a RISK^INDEX^ value which corresponds to an estimated sensitivity, specificity, PPV and NPV with 95% confidence intervals. Lines are the point estimates at each RISK^INDEX^ and shaded regions are the 95% confidence intervals.

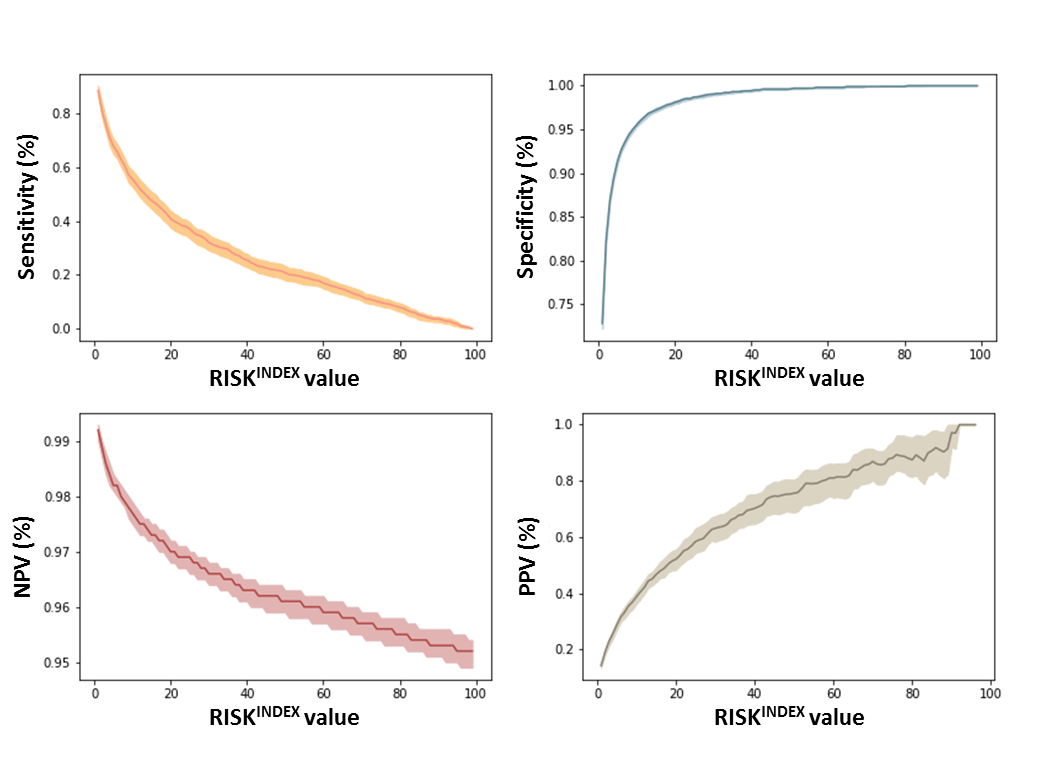

**Supplemental Figure 5. Embedded reference figures for machine learning model in Heerlen.** For each patient in the Zuyderland medical center, location Heerlen the model outputs a RISK^INDEX^ value which corresponds to an estimated sensitivity, specificity, PPV and NPV with 95% confidence intervals. Lines are the point estimates at each RISK^INDEX^ and shaded regions are the 95% confidence intervals.

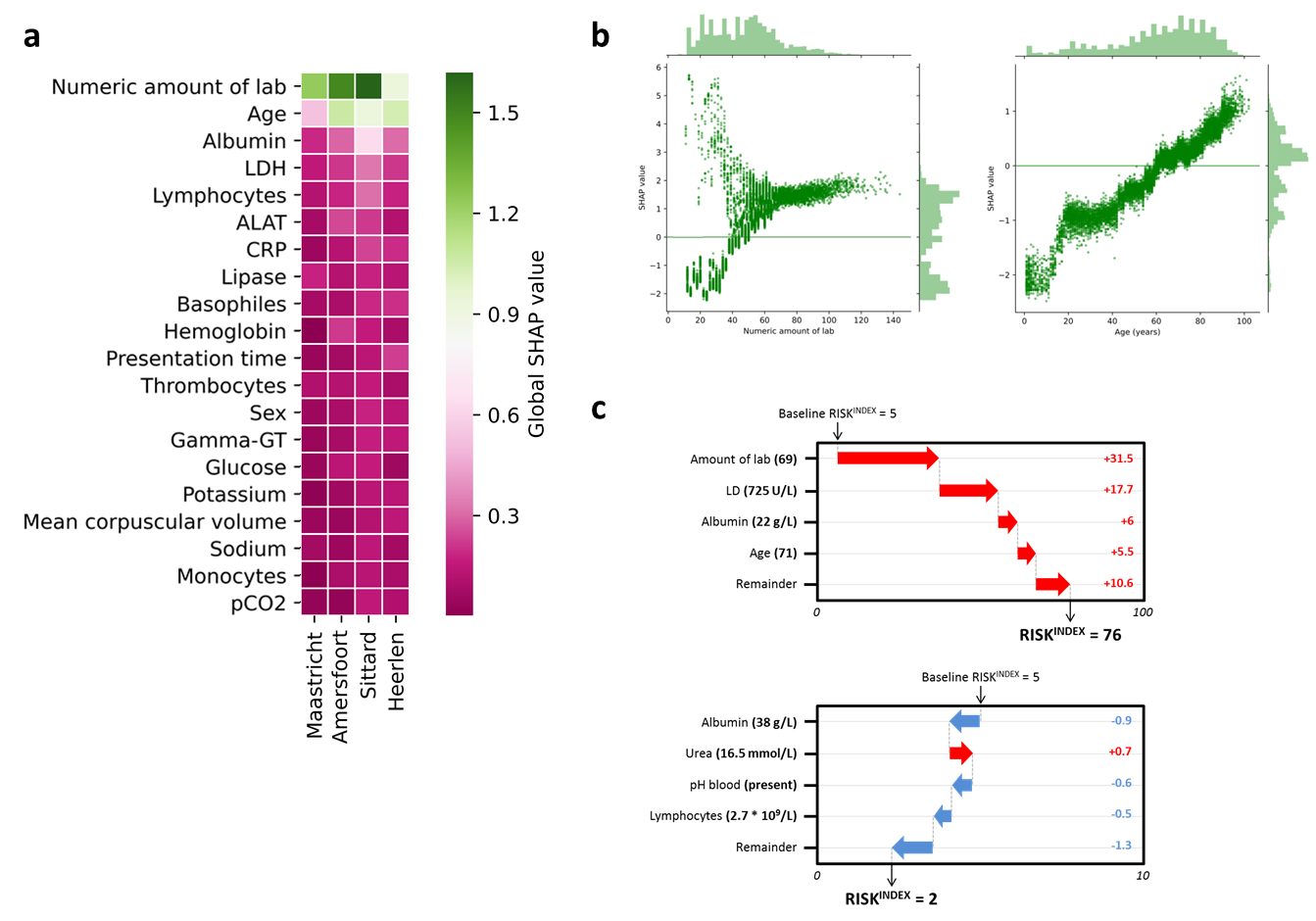

**Supplemental Figure 6. Impact of model parameters.** (A) Heatmap of the global SHAP values for the developed models in each of the centers. Features were ordered from highest to lowest impact averaged over the four models. Green features indicate high impact; purple features indicate lower overall impact. (B) Specific examination of the relationship between the top-2 features (numeric amount of laboratory requests and age) and the SHAP value in the machine learning model of Maastricht. We found that the number of laboratory measurements possesses a ‘double’ relationship with the SHAP value, whilst the age of an individual shows a positive, almost linear relationship with the SHAP value. (C) Illustrative example of how patient characteristics and laboratory results build up to a RISK^INDEX^ for a high- (upper panel) and low-risk (lower panel) individual.

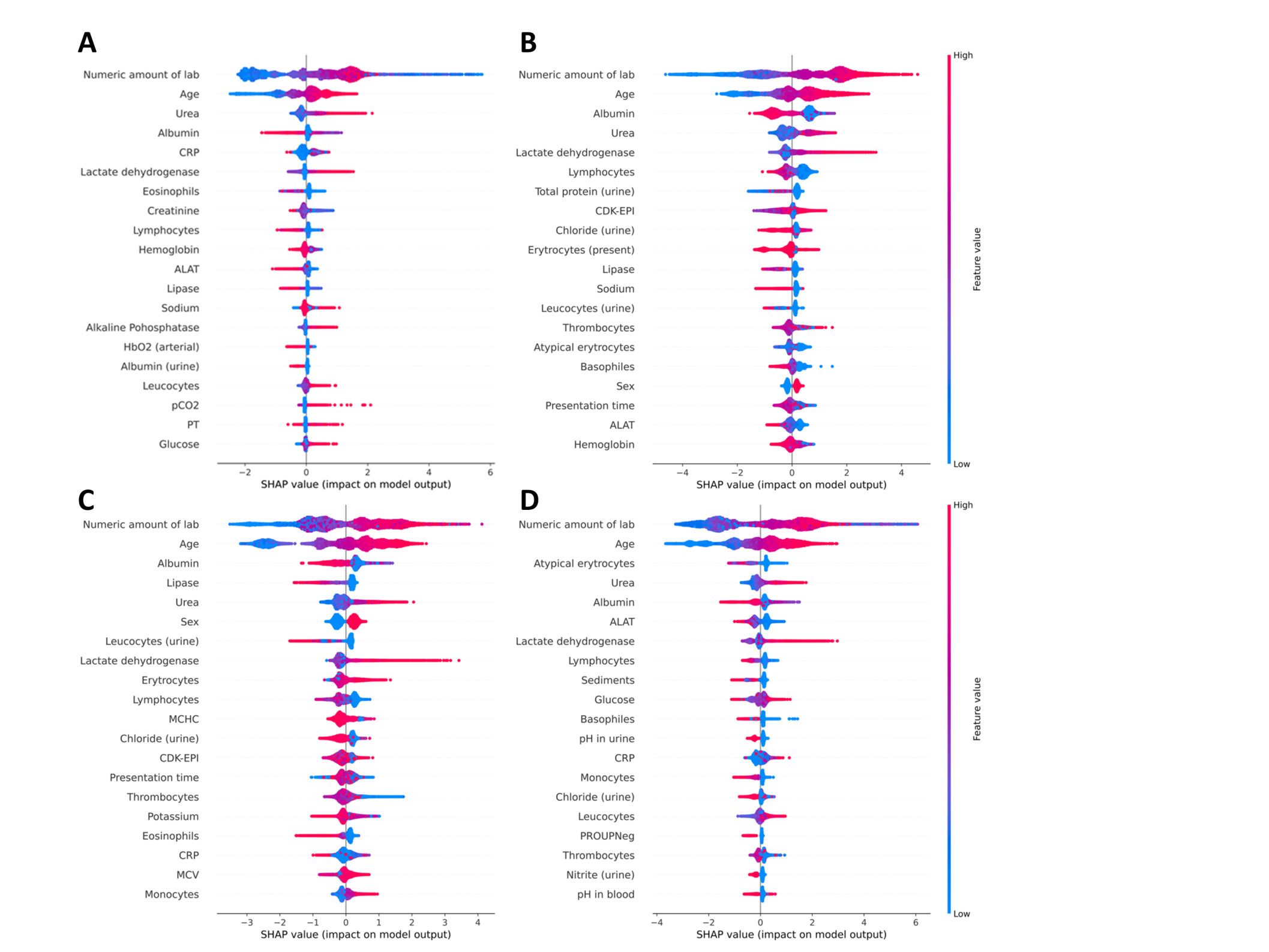

**Supplemental Figure 7. Global SHAP values in each of the centers.** Analysis of feature importance in the LightGBM model developed in Maastricht (A), Sittard (B), Heerlen (C) and Meander (D) using SHAP values. The features are ranked by importance in descending order based on the sum of the SHAP values over all the samples

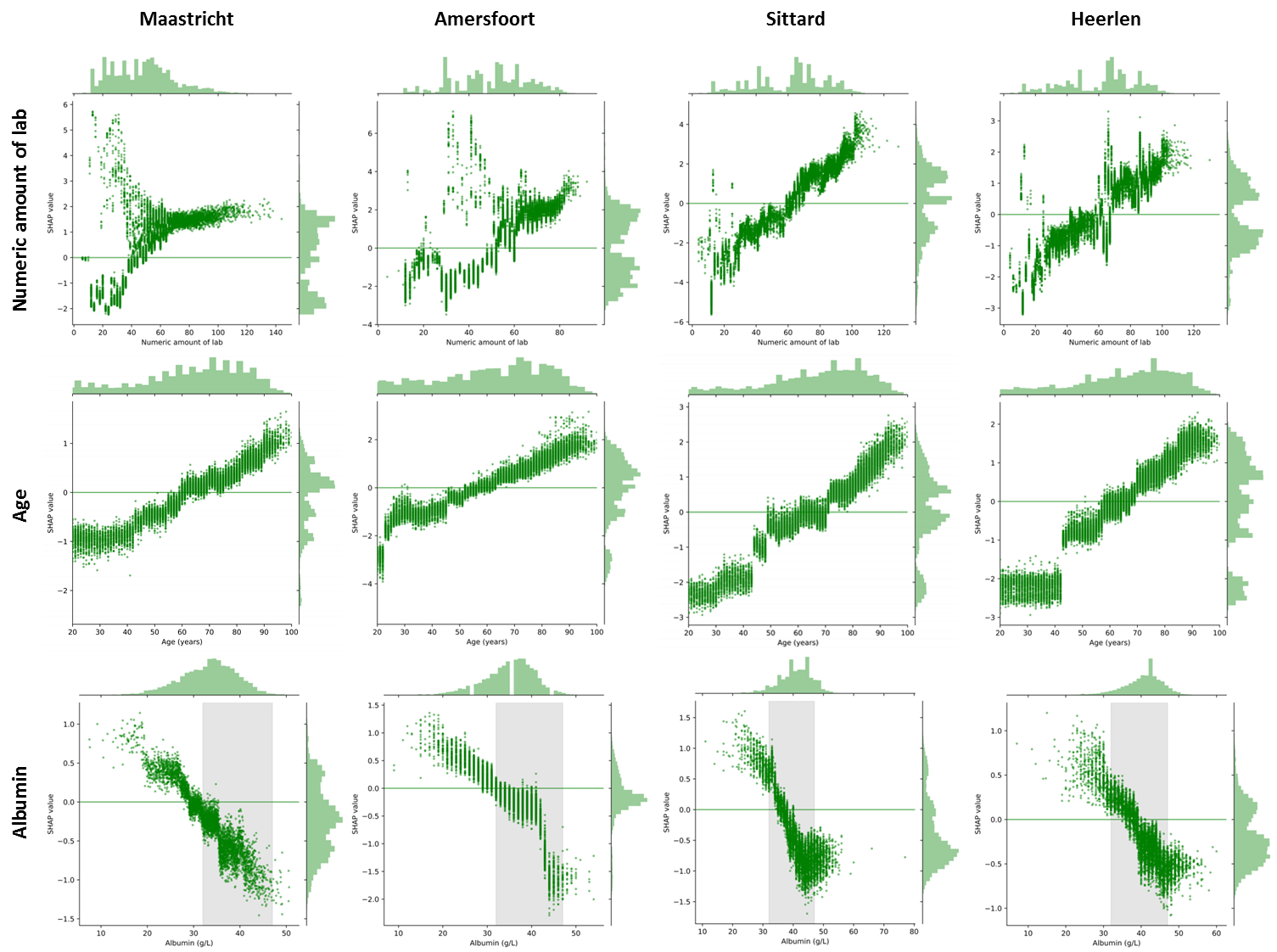

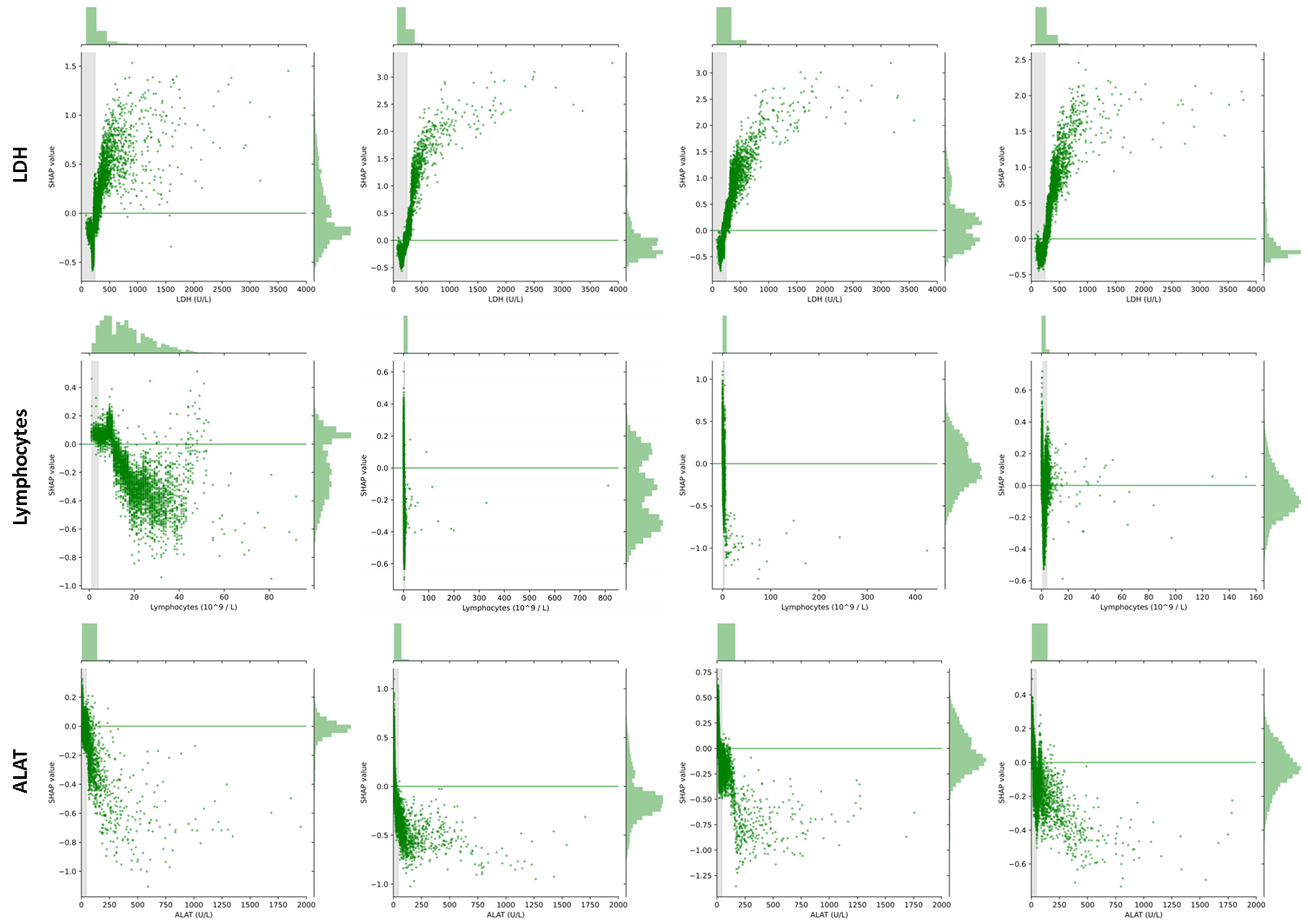

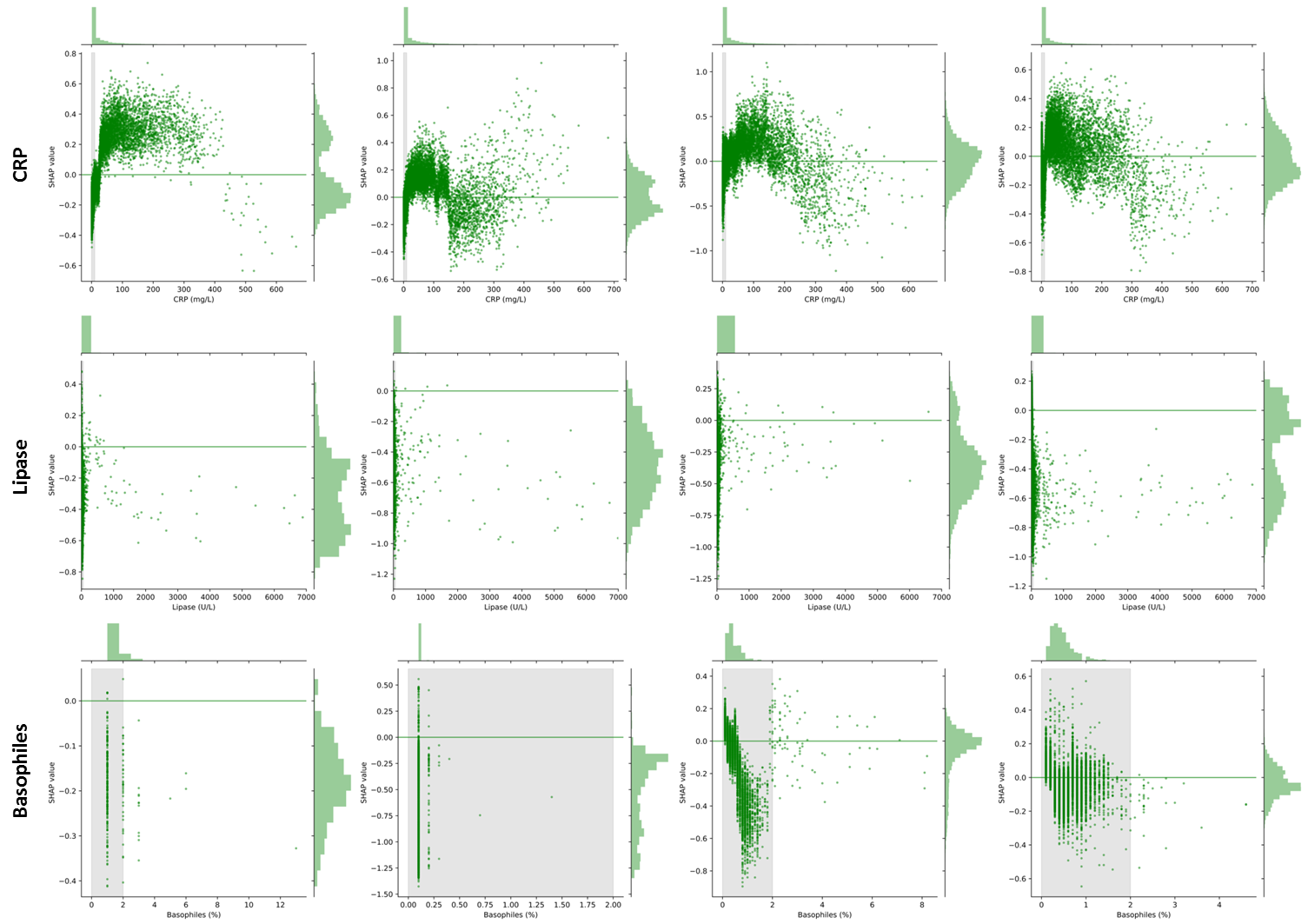

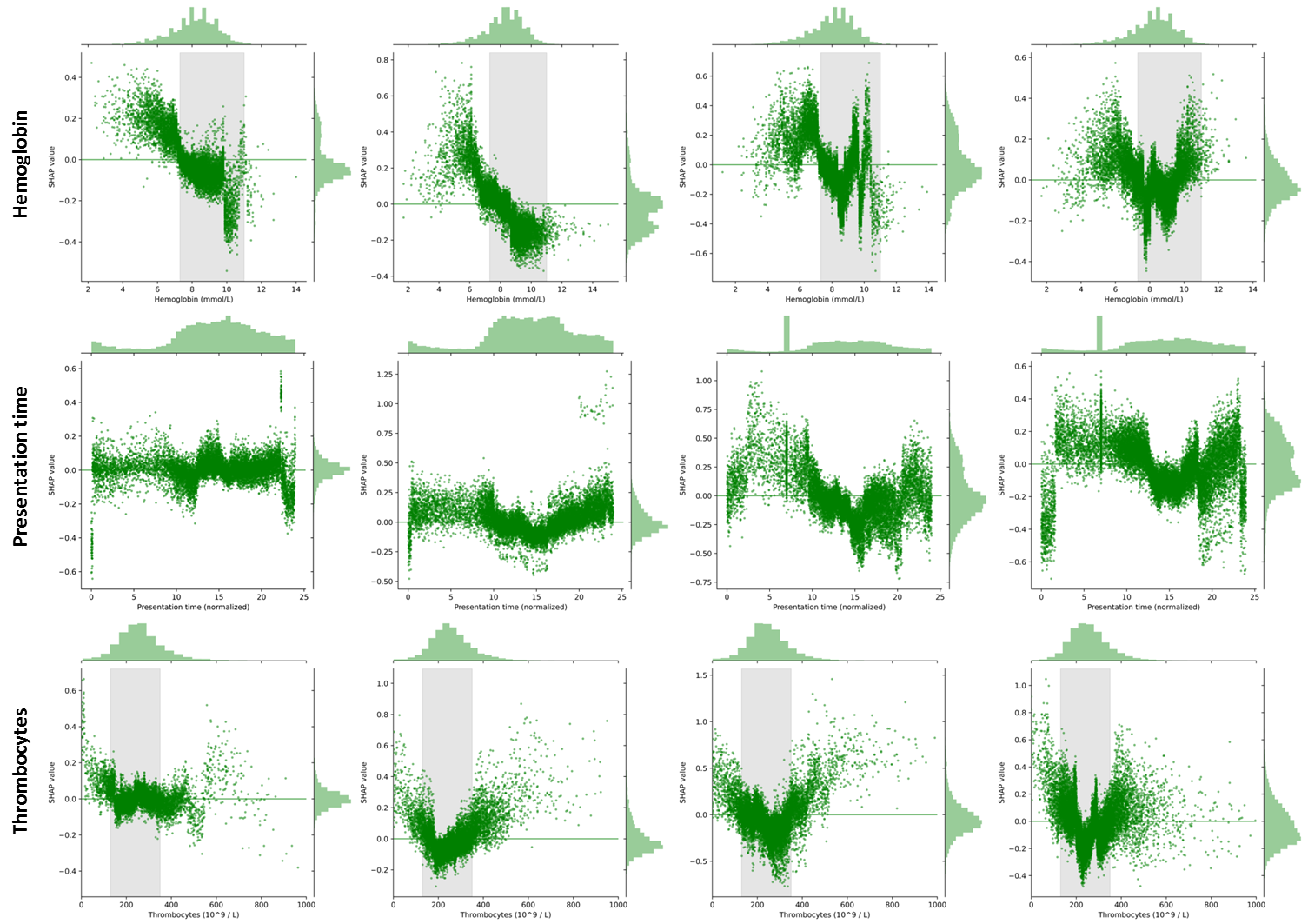

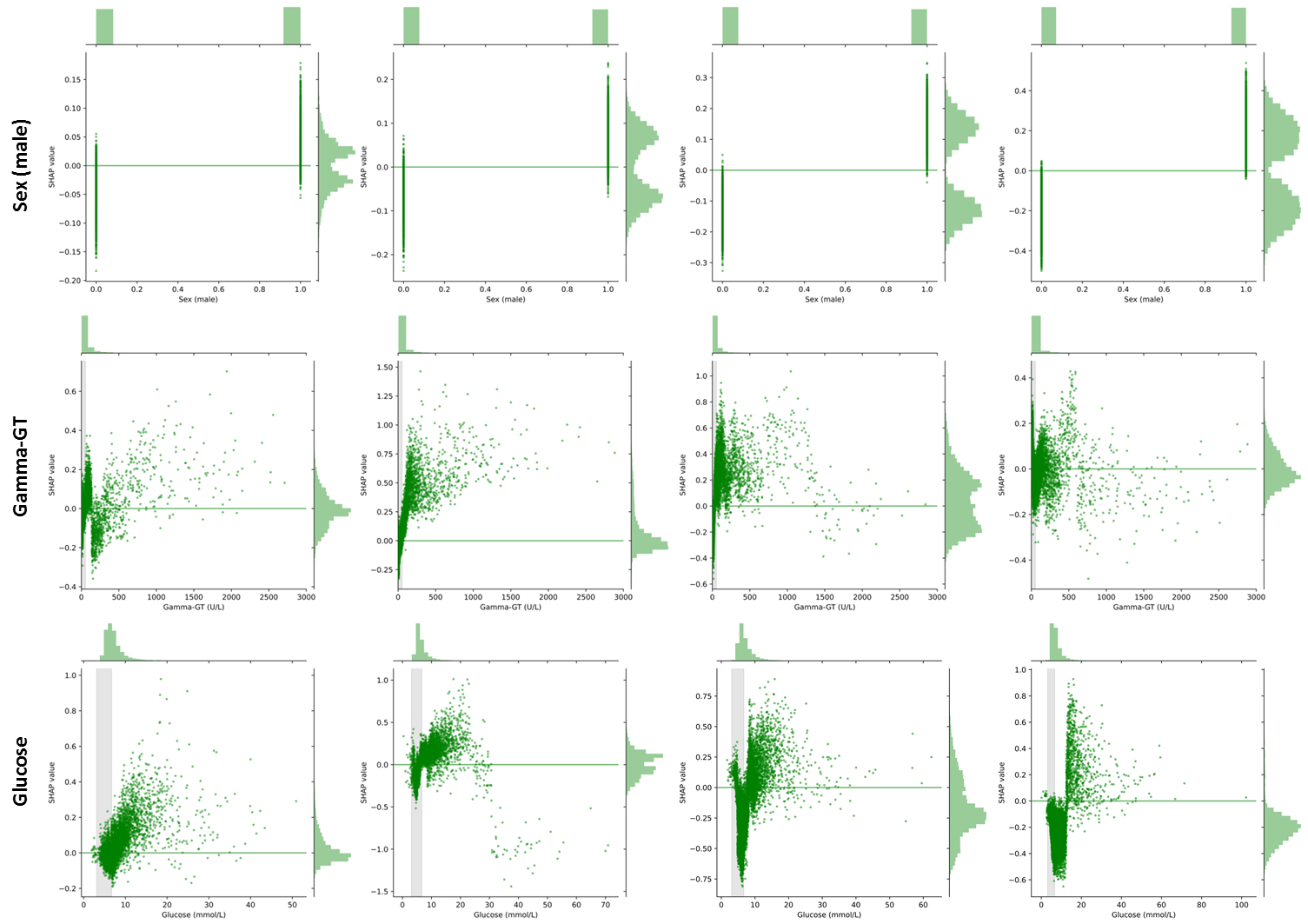

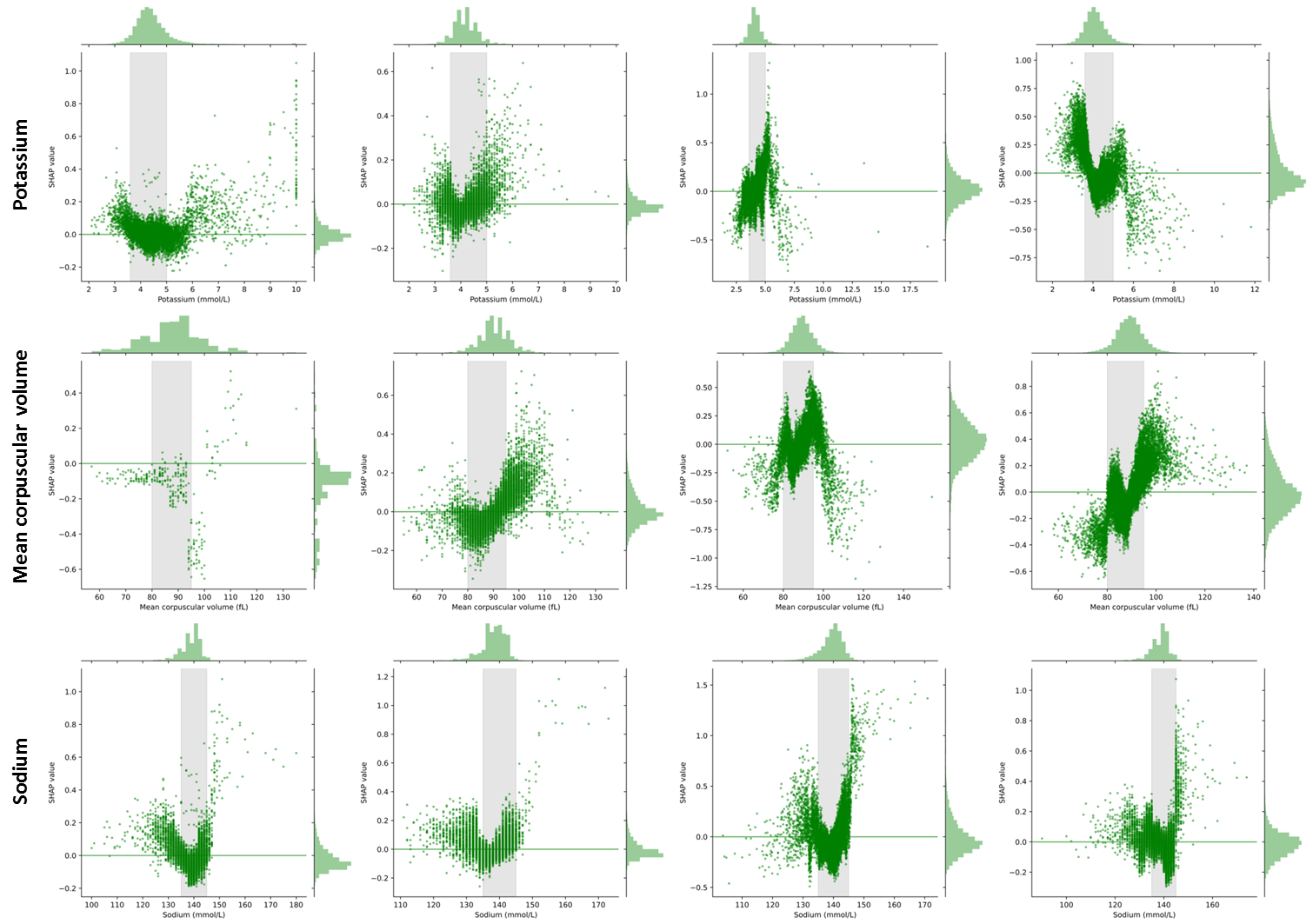

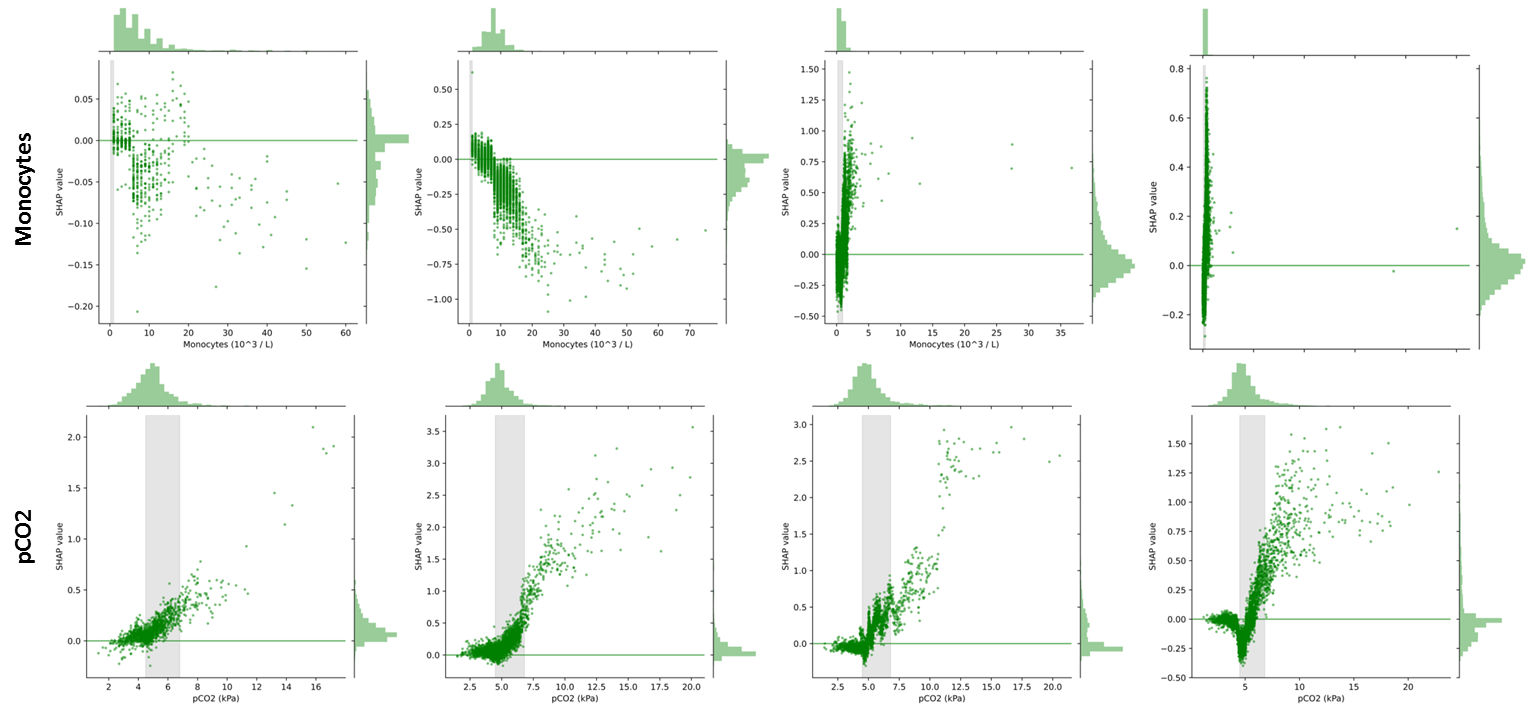

**Supplemental Figure 8. Impact of individual features on the SHAP value.** The relationship of each feature from the top-20 features with the SHAP value was evaluated for each of the centers (left to right: Maastricht, Amersfoort, Sittard and Heerlen). Individual points are depicted on the X-axis with their associated SHAP value on the Y-axis. Reference ranges -if available- for the specific laboratory test are shown as a grey area. Distributions of the values as well as the SHAP values are plotted on the outside borders of each graph.
